## Supplementary Tables and Figure for "Low Haemoglobin Levels are Associated with Reduced Psychomotor and Language Abilities in Young Ugandan Children"

### **Table 1:** Observational studies investigating associations between iron status and neurobehavioural outcomes in African pre-school children.

| **Author, year (country)** | **Sample** | **Study design** | **Age** | **Domain** | **Assessment tool** | **Definition of iron status** | **Results** |
| --- | --- | --- | --- | --- | --- | --- | --- |
| Youssef, 2020 (Egypt) | 226 children | Cross-sectional | 2-6 years | IQ  Language | SBIS  Arabic language test | Not included | No association with IQ or language development. |
| Rothman, 2018 (South Africa) | 750 infants | Cross-sectional | 6 months | Psychomotor | KDI  Parent rating scale | Anaemia: Hb<110 g/L ID: plasma ferritin<12 μg/L  IDA:ID+ Hb < 110 g/L | No association with psychomotor development. |
| Prado, 2017 (Ghana, Malawi, and Burkina Faso) | Mother-child pairs: Ghana (n=1023) and Malawi (n=675)  Child cohorts: Malawi (n=1385) and Burkina Faso (n=1122) | Cohort | 18 months | Motor  Language | CDI  DMC  KDI | Anaemia: Hb<110 g/L | No associations between maternal Hb or iron status during pregnancy and motor or language development at 18 months  Child Hb and iron status at six to nine months were associated with motor and language development at 18 months |
| Mireku, 2015 and 2016* (Benin) | 636 mother-child pairs | Cohort | Maternal iron status and Hb at median  23 (19–26) weeks + cord blood ferritin and Hb at birth and neuroassessment in children at 1 year | Cognitive  Motor  Language | Mullen Scales of Early  Learning | ID: SF<12 μg/L or SF 12-70 μg/L with inflammation: CRP >5 mg/L  Maternal anaemia: Hb < 10 g/L  Child anaemia birth: Hb <140 g/L  IDA: ID + anaemia | An inverted U-shaped association between maternal Hb and gross motor development with Hb 90–110 g/L being optimal for gross motor development.  No association between maternal iron status during pregnancy or cord blood ferritin and Hb levels and cognitive or language development at one year. |
| Gashu, 2016 (Ethiopia) | 541 preschool children | Cross-sectional | 54-60 months | Cognitive | WPPSI-III, the school readiness test. | ID: SF<12μg/l  Anaemia: Hb<110 g/l adjusted for altitude  IDA: ID + Hb<110 g/l | Anaemia was associated with lower mean cognitive scores compared to normal Hb status  No association between ID/ IDA and cognitive development. |
| Angulo-Barroso, 2011 (Ghana, USA, China) | 209 infants (49 from Ghana, 113 from USA and 47 from China) | Multisite cross-sectional | 9-10 months | Motor | Pictorial milestone chart | IDA: Hb < 110 g/L + MCV <74 fl and/or RDW >14%  ID: Hb ≥ 110 g/L + MCV < 74 fl and/or RDW > 14%  IS: Hb ≥ 110 g/L, MCV ≥ 74 fl, and RDW ≤ 14% | IDA and ID were associated with lower motor function compared to sufficient iron status across the three study populations. |
| Olney, 2007  (Zanzibar) | 771 children | Cross-sectional | 5-19 months | Motor | A 20 mutually exclusive item code | IDA: Hb <100 g/L and ZnPP ≥90 mmol/mol heme  ID: ZnPP ≥90 mmol/mol heme  Anaemia: Hb <100 g/L | IDA and ID were associated with lower motor function compared to sufficient iron status. |
| Kariger, 2005  (Zanzibar) | 646 children | Cross-sectional | 6-18 months | Motor (walking and crawling) | Pictorial milestone chart | ID: ZnPP≥ 90 µmol/mol heme  IDA: Hb<100 g/L, ZnPP ≥90 mol/mol heme  IS: Hb≥ 100 g/L, ZnPP <90 µmol/mol heme  Anaemia: Hb<100 g/L | Anaemia and/or ID was associated with lower odds of walking compared to sufficient iron status  No association with crawling. |
| Bouhouch, 2016 (Morocco) | n=455 lead exposed children (110 received iron, 116 received iron+ EDTA, 112 received EDTA, and 117 received placebo for 28 weeks) | RCT | 3 to 14 years | Cognitive | (KABC-II, HVLT) | ID: SF <12 mg/L for children <5 years, SF <15 mg/L for children ≥5 years, or TfR >8.3 mg/L with (CRP ≤5 mg/L, α1-acid glycoprotein ≤51 g/L)  Anaemia: Hb<11.0 g/dL for children <5 years, Hb<11.5 g/dL for children 5–11 years, Hb <12.0 g/dL for | No difference in cognitive scores between children who received iron supplementation or placebo. |
| Baumgartner, 2012  (South Africa) | n=288 (70 received iron+ placebo, 72 placebo + DHA/EPA, 73 iron+ DHA/EPA, and 73 placebo + placebo for 8.5 months) | RCT | 6 to 11 years | Cognitive | (HVLT, KABC) | ID: SF<15 µg/L excluding children with CRP >5 mg/L or ZnPP >70 µmol/mol or TfR >8.3 mg/L  Anaemia: Hb <11.5 g/dL  IDA: anaemia + SF<15 µg/L | Anaemic children who received iron supplementation+ placebo had higher cognitive scores compared to children who received placebo + placebo  No difference in cognitive scores in children who received iron+ DHA/EPA compared to children who received placebo + placebo |
| Stoltzfus, 2001 (Zanzibar) | n=359  (183 received iron and 176 placebo for 12 months) | RCT | 6 to 59 months | Language  Motor | Parents reported motor and language milestones | ID: SF<12 mg/L  Anaemia: Hb<11g/dL  Severe anaemia: Hb<7g/dL | Children who received iron had higher language scores compared to children who received placebo and children with baseline Hb<9g/dL who received iron had higher motor scores compared to children who received placebo. |
| Boivin, 1993 (Zaire) | N=47 (17 children received anthelminthics and iron, 7 only iron, 8 only anthelminthics, and 15 did not receive either intervention for 1 month) | RCT | Mean age for boys=7.7, SD=0.8 years) and for girls =8.0, SD =1.8 years | Cognitive | KABC | Anaemia: Hb<12 g/dL | No difference in cognitive scores between children who received only iron supplementation or placebo. |

CDI, communicative development inventory; DMC, Developmental milestones checklist; Hb, haemoglobin; ID, iron deficiency; IDA, iron deficiency anaemia; IQ, intelligence quotient; IS, iron sufficiency; KDI, Kilifi Developmental Inventory; MCV, mean corpuscular volume; RDW, red blood cell distribution width; SBIS, Stanford Binet intelligence scale version four; WPPSI-III, Wechsler Preschool and Primary Scale of Intelligence; ZnPP, zinc protoporphyrin. * A single mother-child study with two publications separately evaluating the effects of maternal haemoglobin levels and iron status during pregnancy on developmental scores at one year.

### **Table 2:** Measures of motor and cognitive function used at age 15 months

| **Name of test** | **Domain** | **Description of measure** | **Absolute scores (min, max)** |
| --- | --- | --- | --- |
| Fine motor | Motor | Control of small hand-movements was assessed using 25 items such as building a tower with blocks and scribbling with a pen. | 0, 32 |
| Gross Motor | Motor | Control of the limbs assessed using 35 items such as kicking a ball or climbing onto a platform | 0, 35 |
| Self-control | Delay inhibition | This was assessed using two trials in which a biscuit (trial 1) or wrapped gift (trial 2) was presented to the child who was instructed not to open it until the assessor had finished what he/she was doing. Waiting time (in seconds) was recorded to a maximum of 150 seconds. The average time for the two trials was calculated. | 0, 150 |
| The A-not-B task | Contrast inhibition/  working memory | A biscuit was placed in one of two wells as the child watched and both wells were then covered with opaque cups. The board was taken out of sight for 10 seconds during which the child was distracted with a song. The board was then brought back, and the child asked to point to the well with the biscuit. The child was given the biscuit if she or he successfully located it. The location of the biscuit was switched to the other well after two consecutive correct responses. Ten trials were administered, and the number of correct responses recorded. | 0, 10 |
| Language interview | Language | The mother/guardian was interviewed on whether the infant produced common pre-speech items such as vowels (e.g., aa, aa), babble (e.g., ma, ma) or gestures (e.g., waving for ‘‘bye’), spoke definite words, or names of common household objects (up to 11 items). | 0, 22 |
| Recognition of self and others | Socio-cognition | The mother/guardian was interviewed on whether the infant responded to his/her name or distinguished his mother and other familiar people from strangers (12 items). | 0, 24 |
| Self-care |  | The mother/guardian was asked about their child’s behaviours such as how much she/he participated during dressing and feeding (15 items). | 0, 30 |

### **Table 3.** Principal component analysis, loadings on the 3 developmental components at 5 years of age

| **Measure** | **Component 1 (verbal and non-verbal IQ)** | **Component 2 (executive function)** | **Component 3 (motor ability)** |
| --- | --- | --- | --- |
| **Block design** | 0.65 | -0.12 | 0.08 |
| **Picture vocabulary scale** | 0.57 | 0.17 | -0.07 |
| **Verbal fluency** | -0.15 | 0.59 | 0.13 |
| **Picture search** | 0.09 | 0.56 | 0.05 |
| **Wisconsin card sort test** | 0.32 | 0.41 | -0.24 |
| **Coin box** | -0.16 | 0.24 | 0.71 |
| **Balancing on one leg** | 0.31 | -0.24 | 0.63 |

IQ, intelligence quotient. Component 1 (verbal and non-verbal IQ) was strongly positively correlated and with block design and picture vocabulary tests, component 2 (executive function) was positively correlated with verbal fluency, picture search and Wisconsin card sort tests and component 3 (motor ability) was strongly correlated with coinbox and balancing on one leg tests. The component scores are centred on zero with high scores indicating better and low scores worse development.

### **Table 4:** Measures of motor and cognitive function used at age 5 years

| **PCA Components** | **Name of test** | **Domain** | **Description of measure** | **Absolute scores (min, max)** |
| --- | --- | --- | --- | --- |
| Verbal and non-verbal IQ | Block design | Non-verbal IQ | The measure is adapted from the British Ability Scales-third edition. The child is asked to copy and construct items with wooden blocks following a demonstration by the assessor. | 0, 16 |
|  | Picture vocabulary scale | Verbal IQ | The measure is adapted from the Kilifi Vocabulary Test. The child is asked to point out and identify items from 24 black and white picture items familiar to them. | 0, 24 |
| Executive function | Verbal fluency | Working memory | The measure is adapted from the Developmental NEuroPSYchological Assessment. The child is asked to name items including foods and animals as fast as possible in a minute. | a |
|  | Picture search | Selective attention | The measure is adapted from the Sky Search in Tests of Everyday Attention for Children. The child is presented with three A3 sheets each with a target picture on top and about 100 others at the bottom including copies of the target picture. The child is asked to locate as many copies of the target pictures as possible within 10 seconds. | b |
|  | Wisconsin card sort test | Cognitive flexibility | The measure is adapted from Berg’s card sort test. The child is given four playing cards of different suits and a pack of 12 cards and asked to sort the cards by number (block 1) and suit (block 2). | 0,12 |
| Motor function | Coin box | Fine motor function | The measure is adapted from the Kilifi Developmental Inventory. The child is asked to slot coins through a small opening on a coinbox within 20 seconds in two trials. | 0, 20* |
|  | Balancing on one leg | Gross motor function | The measure is adapted from the Movement Assessment Battery for Children. It entails timed attempts (two per leg) of balancing on one leg for one minute. | 0, 60* |

Abbreviations: PCA, principal components analysis; IQ, intelligence quotient; Min, minimum score; Max, maximum score. ^a^ One point is awarded for each correct name and a total score is calculated from the total correct names in a minute. ^b^ A total score is calculated from the number of target pictures identified within 10 seconds. *An average score is calculated after timed attempts of the tests. Block design and picture vocabulary tests loaded heavily on verbal and non-verbal IQ while verbal fluency, picture search, and Wisconsin card sort tests loaded heavily on executive function. Coin box and balancing on one leg tests loaded heavily on motor function. The resulting scores were centred on zero with higher scores representing better and lower scores worse development.

### **Table 5.** Maternal characteristics at enrolment.

| **Variable** | **Analysis for aim 1; maternal Hb during pregnancy, child Hb 12mths and development at 15 months (n=933)** | **Analysis for aim 2; annual Hb and development at 5 years (n=726)** | **Analysis for aim 3; iron status at 2 years and development at 5 years (n=530)** |
| --- | --- | --- | --- |
| Maternal age (years), mean (SD) | 24.2 (5.6) | 24.2 (5.6) | 24.5 (5.6) |
| Maternal education, n/total (%) |  |  |  |
| Primary/none | 494/930 (53.1) | 393/724 (54.3) | 295/528 (55.9) |
| Secondary | 356/930 (38.3) | 273/724 (37.7) | 193/528 (36.6) |
| Tertiary | 80/930 (8.6) | 58/724 (8.0) | 40/428 (7.6) |
| Maternal Parity, n/total (%) |  |  |  |
| 1 | 226/933 (24.2) | 167/726 (23.0) | 111/530 (20.9) |
| 2-4 | 534/933 (57.2) | 419/726 (57.7) | 310/530 (58.5) |
| 5+ | 173/933 (18.5) | 140/726 (19.3) | 109/530 (20.6) |
| Maternal treatment with albendazole, n/total (%) | 475/933 (51.0) | 366/726 (50.4) | 272/530 (51.3) |
| Maternal treatment with praziquantel, n/total (%) | 460/933 (49.3) | 349/726 (48.1) | 250/530 (47.2) |

SD, standard deviation.

### **Table 6:** Distribution of individual developmental measures at 15 months and 5 years of age.

| **Measure** | **N** | **Obtained Median (interquartile range) scores** |
| --- | --- | --- |
| **At 15 months** | | |
| **Fine motor** | 920 | 17 (16-18) |
| **Gross motor** | 936 | 18 (16-19) |
| **Psychomotor (fine motor + gross motor)** | 919 | 35 (32-37) |
| **Language** | 933 | 15 (12-19) |
| **Recognition of self and others** | 933 | 10 (10-10) |
| **Self-care** | 933 | 18 (14-20) |
| **Social cognition (recognition of self and others + self-care)** | 933 | 27 (24-30) |
| **Self-control** | 805 | 1.5 (1-3.5) |
| **A not B task** | 758 | 4 (2-6) |
| **Executive function**  **(A not B + self-control)** | 689 | 7 (5-9) |
| **At 5 years** | | |
| **Block design** | 804 | 9 (6-11) |
| **Picture vocabulary scale** | 811 | 18 (16-20) |
| **Verbal fluency** | 760 | 14 (8-20) |
| **Picture search** | 811 | 4 (3-5) |
| **Wisconsin card sort test** | 807 | 6 (3-10) |
| **Coin box** | 807 | 10 (9-11) |
| **Balancing on one leg** | 802 | 12 (7-20.3) |

### **Table 7:** Univariable and multivariable linear regression results for the associations between participant characteristics and developmental scores at 15 months.

| **Variables** | **n** | **Executive function**  **β (95% CI)** | **P value** | **n** | **Psychomotor function**  **β (95% CI)** | **P value** | **n** | **Social cognition**  **β (95% CI)** | **P value** | **n** | **Language**  **β (95% CI)** | **P value** |
| --- | --- | --- | --- | --- | --- | --- | --- | --- | --- | --- | --- | --- |
| **Univariable linear regression results for the associations with developmental scores at 15 months** | | | | | | | | | | | | |
| **Child characteristics** |  |  |  |  |  |  |  |  |  |  |  |  |
| Age at neuroassessment | 687 | 0.01 (-0.13, 0.14) | 0.94 | 916 | 0.14 (0.02, 0.26) | 0.02 | 934 | -0.02 (-0.21, 0.17) | 0.84 | 934 | 0.09 (-0.03, 0.21) | 0.14 |
| Sex (female) | 688 | -0.04 (-0.19, 0.11) | 0.59 | 918 | -0.16 (-0.29, -0.03) | 0.01 | 936 | 0.18 (-0.03, 0.38) | 0.09 | 936 | 0.02 (-0.11, 0.15) | 0.72 |
| Malaria parasitemia at 1^st^ annual visit (Positive) | 686 | 0.25 (-0.11, 0.60) | 0.17 | 915 | -0.39 (-0.69, -0.09) | 0.01 | 933 | -0.39 (-0.87, 0.08) | 0.10 | 933 | -0.16 (-0.46, 0.13) | 0.28 |
| Clinical malaria episodes at 1 year of age | 688 | -0.04 (-0.16, 0.07) | 0.47 | 918 | -0.11 (-0.21, -0.02) | 0.02 | 936 | -0.04 (-0.13, 0.06) | 0.43 | 936 | 0.004 (-0.09, 0.10) | 0.94 |
| Any worm infections at 1^st^ annual visit | 659 | -0.04 (-0.52, 0.44) | 0.88 | 889 | 0.10 (-0.30, 0.51) | 0.62 | 905 | -0.16 (-0.82, 0.50) | 0.64 | 905 | -0.32 (-0.74, 0.09) | 0.13 |
| Child treatment with albendazole (placebo) | 687 | -0.03 (-0.18, 0.11) | 0.66 | 916 | 0.01 (-0.12, 0.14) | 0.90 | 934 | -0.11 (-0.31, 0.10) | 0.30 | 934 | -0.07 (-0.20, 0.06) | 0.28 |
| **Nutritional status** |  |  |  |  |  |  |  |  |  |  |  |  |
| Stunting | 680 | 0.10 (-0.12, 0.32) | 0.39 | 907 | -0.53 (-0.72, -0.35) | <0.001 | 925 | -0.31 (-0.60, -0.01) | 0.04 | 925 | -0.28 (-0.47, -0.09) | 0.003 |
| Underweight | 688 | -0.17 (-0.43, 0.09) | 0.21 | 918 | -0.55 (-0.77, -0.32) | <0.001 | 936 | -0.34 (-0.70, 0.02) | 0.07 | 936 | -0.23 (-0.45, -0.002) | 0.05 |
| Wasting | 680 | 0.01 (-0.38, 0.37) | 0.98 | 907 | -0.55 (-0.87, -0.24) | 0.001 | 925 | -0.12 (-0.63, 0.39) | 0.65 | 925 | -0.09 (-0.41, 0.24) | 0.60 |
| **Maternal characteristics at enrolment** |  |  |  |  |  |  |  |  |  |  |  |  |
| Maternal age (years) | 689 |  |  | 919 |  |  | 933 |  |  | 933 |  |  |
| 14-24 |  | Reference |  |  | Reference |  |  | Reference |  |  | Reference |  |
| 25-34 |  | -0.04 (-0.20, 0.12) |  |  | 0.10 (-0.04, 0.24) |  |  | 0.18 (-0.04, 0.39) |  |  | 0.10 (-0.03, 0.24) |  |
| 35+ |  | -0.16 (-0.47, 0.14) | 0.31* |  | 0.18 (-0.08, 0.45) | 0.07* |  | 0.16 (-0.27, 0.59) | 0.13* |  | -0.06 (-0.32, 0.21) | 0.49* |
| Maternal education | 686 |  |  | 916 |  |  | 934 |  |  | 934 |  |  |
| Primary/none |  | 0.16 (-0.11, 0.43) |  |  | -0.29 (-0.53, -0.06) |  |  | 0.18 (-0.20, 0.56) |  |  | -0.15 (-0.39, 0.08) |  |
| Secondary |  | 0.19 (-0.09, 0.46) |  |  | -0.09 (-0.33, 0.16) |  |  | 0.31 (-0.08, 0.70) |  |  | 0.07 (-0.16, 0.32) |  |
| Tertiary |  | Reference | 0.50* |  | Reference | 0.001* |  | Reference | 0.99* |  | Reference | 0.01* |
| Parity | 689 |  |  | 919 |  |  | 933 |  |  | 933 |  |  |
| 1 |  | Reference |  |  | Reference |  |  | Reference |  |  | Reference |  |
| 2-4 |  | -0.01 (-0.19, 0.17) |  |  | -0.02 (-0.18, 0.13) |  |  | 0.09 (-0.15, 0.34) |  |  | -0.03 (-0.19, 0.12) |  |
| 5+ |  | 0.01 (-0.22, 0.23) | 0.99* |  | -0.05 (-0.25, 0.15) | 0.63* |  | 0.14 (-0.17, 0.45) | 0.37* |  | -0.03 (-0.23, 0.17) | 0.77* |
| Albendazole treatment in pregnancy (placebo) | 689 | -0.05 (-0.20, 0.09) | 0.48 | 919 | -0.05 (-0.18, 0.08) | 0.43 | 933 | -0.10 (-0.30, 0.10) | 0.34 | 933 | 0.02 (-0.10, 0.15) | 0.71 |
| Praziquantel treatment in pregnancy (placebo) | 689 | -0.15 (-0.29, -0.002) | 0.05 | 919 | 0.03 (-0.10, 0.16) | 0.62 | 933 | -0.10 (-0.30, 0.11) | 0.36 | 933 | 0.05 (-0.08, 0.18) | 0.46 |
| Household social economic status | 675 |  |  | 902 |  |  | 919 |  |  | 919 |  |  |
| 1 (lowest) |  | 0.09 (-0.34, 0.53) |  |  | -0.05 (-0.43, 0.33) |  |  | -0.22 (-0.83, 0.40) |  |  | -0.37 (-0.76, 0.01) |  |
| 2 |  | 0.08 (-0.32, 0.49) |  |  | -0.31 (-0.67, 0.05) |  |  | -0.01 (-0.58, 0.56) |  |  | -0.20 (-0.56, 0.16) |  |
| 3 |  | 0.09 (-0.24, 0.42) |  |  | -0.31 (-0.60, -0.02) |  |  | -0.11 (-0.57, 0.36) |  |  | -0.27 (-0.57, 0.02) |  |
| 4 |  | 0.11 (-0.19, 0.50) |  |  | -0.05 (-0.34, 0.25) |  |  | 0.26 (-0.21, 0.74) |  |  | -0.08 (-0.37, 0.22) |  |
| 5 |  | 0.16 (-0.19, 0.50) |  |  | -0.09 (-0.39, 0.21) |  |  | 0.06 (-0.42, 0.55) |  |  | -0.07 (-0.38, 0.23) |  |
| 6 (highest) |  | Reference | 0.87* |  | Reference | 0.04* |  | Reference | 0.13* |  | Reference | 0.01* |
| **Multivariable linear regression results for the associations with developmental scores at 15 months**** | | | | | | | | | | | | |
| **Child characteristics** |  |  |  |  |  |  |  |  |  |  |  |  |
| Age at neuroassessment | 624 | 0.04 (-0.10, 0.19) | 0.57 | 835 | 0.16 (0.04, 0.28) | 0.01 | 846 | -0.01 (-0.22, 0.19) | 0.91 | 850 | 0.10 (-0.03, 0.22) | 0.14 |
| Sex (female) | 621 | -0.07 (-0.23, 0.09) | 0.41 | 831 | -0.25 (-0.38, -0.12) | <0.001 | 846 | 0.15 (-0.07, 0.37) | 0.19 | 846 | -0.02 (-0.15, 0.12) | 0.82 |
| Malaria parasitemia at 1^st^ annual visit (Positive) | 621 | 0.17 (-0.22, 0.56) | 0.39 | 831 | -0.21 (-0.52, 0.11) | 0.20 | 846 | -0.37 (-0.88, 0.15) | 0.16 | 846 | -0.04 (-0.36, 0.28) | 0.80 |
| Clinical malaria episodes at 1 year of age | 624 | -0.05 (-0.17, 0.08) | 0.47 | 835 | -0.06 (-0.16, 0.03) | 0.18 | 850 | -0.02 (-0.12, 0.08) | 0.75 | 850 | 0.05 (-0.05, 0.15) | 0.33 |
| **Nutritional status** |  |  |  |  |  |  |  |  |  |  |  |  |
| Stunting | 621 | 0.07 (-0.17, 0.31) | 0.57 | 831 | -0.51 (-0.70, -0.31) | <0.001 | 846 | -0.32 (-0.63, 0.004) | 0.05 | 846 | -0.27 (-0.47, 0.08) | 0.01 |
| Underweight | 629 | -0.22 (-0.50, 0.07) | 0.14 | 842 | -0.48 (-0.72, -0.25) | <0.001 | 857 | -0.31 (-0.69, 0.08) | 0.12 | 857 | -0.14 (-0.38, 0.10) | 0.26 |
| Wasting | 621 | 0.003 (-0.41, 0.41) | 0.99 | 831 | -0.49 (-0.82, -0.16) | 0.004 | 846 | -0.04 (-0.59, 0.52) | 0.89 | 846 | -0.02 (-0.36, 0.33) | 0.93 |
| **Maternal characteristics at enrolment** |  |  |  |  |  |  |  |  |  |  |  |  |
| Maternal age (years) | 621 |  |  | 831 |  |  | 846 |  |  | 846 |  |  |
| 14-24 |  | Reference |  |  | Reference |  |  | Reference |  |  | Reference |  |
| 25-34 |  | -0.01 (-0.18, 0.16) |  |  | 0.06 (-0.07, 0.20) |  |  | 0.16 (-0.08, 0.39) |  |  | 0.08 (-0.07, 0.22) |  |
| 35+ |  | -0.25 (-0.58, 0.08) | 0.31* |  | 0.24 (-0.04, 0.53) | 0.09* |  | 0.18 (-0.29, 0.65) | 0.18* |  | 0.01 (-0.29, 0.30) | 0.49* |
| Maternal education | 621 |  |  | 831 |  |  | 846 |  |  | 846 |  |  |
| Primary/none |  | 0.15 (-0.13, 0.44) |  |  | -0.21 (-0.44, 0.03) |  |  | 0.24 (-0.16, 0.64) |  |  | -0.07 (-0.32, 0.18) |  |
| Secondary |  | 0.18 (-0.11, 0.47) |  |  | -0.03 (-0.27, 0.21) |  |  | 0.36 (-0.04, 0.76) |  |  | 0.12 (-0.13, 0.37) |  |
| Tertiary |  | Reference | 0.57* |  | Reference | 0.01* |  | Reference | 0.76* |  | Reference | 0.08* |
| Praziquantel treatment in pregnancy (placebo) | 621 | -0.06 (-0.22, -0.09) | 0.45 | 831 | 0.04 (-0.09, 0.17) | 0.58 | 846 | -0.11 (-0.32, 0.11) | 0.33 | 846 | 0.07 (-0.06, 0.21) | 0.29 |
| Household social economic status | 621 |  |  | 831 |  |  | 846 |  |  | 846 |  |  |
| 1 (lowest) |  | 0.08 (-0.38, 0.54) |  |  | 0.07 (-0.32, 0.45) |  |  | -0.35 (-1.01, 0.30) |  |  | -0.36 (-0.77, 0.04) |  |
| 2 |  | 0.13 (-0.31, 0.57) |  |  | -0.24 (-0.61, 0.13) |  |  | -0.14 (-0.76, 0.48) |  |  | -0.16 (-0.55, 0.23) |  |
| 3 |  | 0.08 (-0.28, 0.44) |  |  | -0.19 (-0.49, 0.11) |  |  | -0.15 (-0.66, 0.35) |  |  | -0.23 (-0.55, 0.08) |  |
| 4 |  | 0.07 (-0.28, 0.43) |  |  | 0.01 (-0.29, 0.31) |  |  | 0.19 (-0.32, 0.69) |  |  | -0.07 (-0.38, 0.25) |  |
| 5 |  | 0.14 (-0.23, 0.50) |  |  | -0.09 (-0.39, 0.22) |  |  | -0.11 (-0.62, 0.41) |  |  | -0.11 (-0.43, 0.21) |  |
| 6 (highest) |  | Reference | 0.97* |  | Reference | 0.35* |  | Reference | 0.17* |  | Reference | 0.03* |

CI, confidence interval. Stunting, underweight and wasting were defined as height-for-age Z scores <-2 SD, weight-for-age Z scores <-2 SD and weight-for-height Z scores <-2 SD respectively. *P value for linear trend. **Multivariable analyses were only conducted for variables that were associated with developmental scores in univariable analyses. Multivariable models were adjusted for age, sex, haemoglobin levels, stunting, socioeconomic status, maternal education, helminthic infections and malaria parasitaemia.

### **Table 8:** Univariable and multivariable linear regression results for associations between participant characteristics and developmental scores at 5 years.

| **Variables** | **n** | **Verbal/nonverbal IQ**  **β (95% CI)** | **P value** | **Executive function**  **β (95% CI)** | **P value** | **Motor ability**  **β (95% CI)** | **P value** |
| --- | --- | --- | --- | --- | --- | --- | --- |
| **Univariable linear regression results for the associations with developmental scores at 5 years** | | | | | | | |
| **Child characteristics** |  |  |  |  |  |  |  |
| Age at iron measurement | 530 | -0.05 (-0.19, 0.09) | 0.49 | -0.08 (-0.21, 0.06) | 0.25 | -0.02 (-0.13, 0.09) | 0.73 |
| Sex (female) | 724 | 0.34 (0.15, 0.54) | 0.001 | -0.05 (-0.24, 0.13) | 0.57 | 0.18 (0.02, 0.33) | 0.03 |
| Malaria parasitaemia during 1-5 years of follow-up | 726 | -0.36 (-0.65, -0.08) | 0.01 | -0.33 (-0.59, -0.06) | 0.01 | -0.30 (-0.52, -0.07) | 0.01 |
| Clinical malaria episodes during 1-5 years of follow-up | 722 | -0,08 (-0.19, 0.03) | 0.15 | -0.05 (-0.16, 0.05) | 0.31 | -0.04 (-0.13, 0.05) | 0.38 |
| Any worm infections during 1-5 years of follow-up | 726 | -0.25 (-0.50, 0.004) | 0.05 | -0.32 (-0.56, -0.08) | 0.01 | -0.11 (-0.31, 0.09) | 0.28 |
| Child treatment with albendazole | 724 | -0.08 (-0.28, 0.12) | 0.43 | 0.06 (-0.13, 0.24) | 0.56 | -0.08 (-0.24, 0.07) | 0.29 |
| **Child nutritional status at 5 years** |  |  |  |  |  |  |  |
| Stunting | 710 | -0.38 (-0.61, -0.16) | 0.001 | -0.73 (-0.94, -0.52) | <0.001 | -0.13 (-0.31, 0.05) | 0.15 |
| Underweight | 714 | -0.40 (-0.73, -0.07) | 0.02 | -0.61 (-0.93, -0.30) | <0.001 | -0.29 (-0.55, -0.02) | 0.04 |
| Wasting | 713 | -0.06 (-0.44, 0.32) | 0.75 | -0.12 (-0.48, 0.24) | 0.51 | -0.11 (-0.42, 0.19) | 0.47 |
| **Maternal characteristics at enrolment** |  |  |  |  |  |  |  |
| Maternal age (years) | 726 |  |  |  |  |  |  |
| 14-24 |  | Reference |  | Reference |  | Reference |  |
| 25-34 |  | 0.19 (-0.02, 0.41) |  | 0.12 (-0.08, 0.32) |  | -0.03 (-0.20, 0.14) |  |
| 35+ |  | -0.13 (-0.52, 0.27) | 0.50* | -0.07 (-0.45, 0.30) | 0.67* | -0.12 (-0.43, 0.20) | 0.46* |
| Maternal education | 724 |  |  |  |  |  |  |
| Primary/none |  | -0.66 (-1.03, -0.29) |  | -0.80 (-1.14, -0.45) |  | 0.05 (-0.25, 0.35) |  |
| Secondary |  | -0.35 (-0.74, 0.03) |  | -0.40 (-0.75, -0.04) |  | 0.10 (-0.20, 0.41) |  |
| Tertiary |  | Reference | <0.001* | Reference | <0.001* | Reference | 0.86* |
| Parity | 726 |  |  |  |  |  |  |
| 1 |  | Reference |  | Reference |  | Reference |  |
| 2-4 |  | 0.09 (-0.16, 0.33) |  | -0.04 (-0.27, 0.19) |  | 0.11 (-0.08, 0.31) |  |
| 5+ |  | -0.22 (-0.52, 0.08) | 0.19* | -0.22 (-0.51, 0.07) | 0.15* | 0.08 (-0.17, 0.32) | 0.49* |
| Albendazole treatment in pregnancy (placebo) | 726 | 0.01 (-0.19, 0.21) | 0.94 | 0.01 (-0.17, 0.20) | 0.88 | 0.10 (-0.06, 0.26) | 0.21 |
| Praziquantel treatment in pregnancy (placebo) | 726 | 0.04 (-0.16, 0.24) | 0.69 | 0.06 (-0.12, 0.25) | 0.41 | 0.07 (-0.09, 0.23) | 0.37 |
| Household social economic status | 713 |  |  |  |  |  |  |
| 1 (lowest) |  | -0.37 (-0.90, 0.17) |  | -0.68 (-1.18, -0.17) |  | -0.05 (-0.49, 0.38) |  |
| 2 |  | -0.01 (-0.54, 0.53) |  | -0.26 (-0.77, 0.25) |  | -0.08 (-0.51, 0.36) |  |
| 3 |  | -0.16 (-0.57, 0.25) |  | -0.26 (-0.65, 0.13) |  | 0.16 (-0.17, 0.49) |  |
| 4 |  | -0.02 (-0.44, 0.39) |  | -0.06 (-0.45, 0.33) |  | 0.20 (-0.13, 0.53) |  |
| 5 |  | 0.37 (-0.06, 0.80) |  | 0.32 (-0.09, 0.72) |  | 0.16 (-0.19, 0.51) |  |
| 6 (highest) |  | Reference | 0.002* | Reference | <0.001* | Reference | 0.43* |
| **Multivariable linear regression results for the associations with developmental scores at 5 years**** | | | | | | | |
| **Child characteristics** |  |  |  |  |  |  |  |
| Sex (female) | 471 | 0.48 (0.25, 0.71) | <0.001 | 0.02 (-0.20, 0.24) | 0.83 | 0.29 (0.10, 0.48) | 0.003 |
| Malaria parasitemia during 1-5 years of follow-up | 471 | -0.46 (-0.80, -0.11) | 0.01 | -0.18 (-0.51, 0.15) | 0.28 | -0.20 (-0.49, 0.09) | 0.17 |
| Any worm infections during 1-5 years of follow-up | 471 | 0.18 (-0.13, 0.48) | 0.26 | 0.02 (-0.27, 0.31) | 0.90 | 0.06 (-0.19, 0.31) | 0.65 |
| **Nutritional status at 5 years** |  |  |  |  |  |  |  |
| Stunting | 471 | -0.07 (-0.33, 0.19) | 0.62 | -0.53 (-0.79, -0.28) | <0.001 | -0.03 (-0.26, 0.19) | 0.76 |
| Underweight | 474 | -0.18 (-0.55, 0.19) | 0.34 | -0.35 (-0.71, 0.01) | 0.06 | -0.17 (-0.48, 0.13) | 0.27 |
| **Maternal characteristics at enrolment** |  |  |  |  |  |  |  |
| Maternal education | 471 |  |  |  |  |  |  |
| Primary/none |  | -0.49 (-0.95, -0.03) |  | -0.47 (-0.91, -0.03) |  | 0.24 (-0.14, 0.62) |  |
| Secondary |  | -0.02 (-0.49, 0.44) |  | -0.09 (-0.54, -0.35) |  | 0.30 (-0.09, 0.68) |  |
| Tertiary |  | Reference | <0.001* | Reference | 0.001* | Reference | 0.61* |
| Household social economic status | 471 |  |  |  |  |  |  |
| 1 (lowest) |  | -0.41 (-1.05, 0.22) |  | -0.37 (-0.98, 0.24) |  | 0.08 (-0.45, 0.61) |  |
| 2 |  | 0.02 (-0.57, 0.62) |  | -0.03 (-0.60, 0.54) |  | 0.08 (-0.42, 0.57) |  |
| 3 |  | -0.24 (-0.71, 0.23) |  | -0.09 (-0.54, 0.36) |  | 0.26 (-0.14, 0.65) |  |
| 4 |  | -0.22 (-0.70, 0.25) |  | 0.03 (-0.42, 0.49) |  | 0.19 (-0.20, 0.59) |  |
| 5 |  | 0.09 (-0.39, 0.59) |  | 0.29 (-0.17, 0.76) |  | 0.16 (-0.25, 0.57) |  |
| 6 (highest) |  | Reference | 0.09* | Reference | 0.02* | Reference | 0.72* |

CI, confidence interval. Stunting, underweight and wasting were defined as height-for-age Z scores <-2 SD, weight-for-age Z scores <-2 SD and weight-for-height Z scores <-2 SD respectively. *P value for linear trend. ** Multivariable analyses only included variables that were associated with developmental scores in the univariable analyses. Multivariable models were adjusted for age, sex, any moderate anaemia event, iron deficiency, stunting, socioeconomic status, maternal education, helminthic infections and malaria parasitaemia.

### **Table 9.** Univariable and multivariable linear regression results for associations between maternal haemoglobin levels and anaemia during pregnancy and developmental scores at 5 years.

| **Developmental domain** | **n** | **Univariable model   β (95% CI)** | **P value** | **n** | **Multivariable model* β (95% CI)** | **P value** | |
| --- | --- | --- | --- | --- | --- | --- | --- |
| **Maternal haemoglobin levels and developmental scores at 5 years** | | | | | | | |
| **Verbal and non-verbal IQ** | 724 | 0.05 (-0.02, 0.12) | 0.14 | 693 | 0.04 (-0.03, 0.11) | 0.29 | |
| **Executive function** | 724 | 0.06 (-0.004, 0.13) | 0.07 | 693 | 0.05 (-0.01, 0.12) | 0.10 | |
| **Motor ability** | 724 | -0.02 (-0.08, 0.03) | 0.42 | 693 | -0.03 (-0.09, 0.02) | 0.25 | |
| **Mild maternal anaemia and developmental scores at 5 years** | | | | | | | |
| **Verbal and non-verbal IQ** | 724 | -0.09 (-0.30, 0.12) | 0.39 | 693 | -0.06 (-0.26, 0.15) | | 0.58 |
| **Executive function** | 724 | -0.12 (-0.32, 0.07) | 0.22 | 693 | -0.12 (-0.31, 0.07) | | 0.21 |
| **Motor ability** | 724 | 0.08 (-0.09, 0.24) | 0.37 | 693 | 0.09 (-0.08, 0.26) | | 0.29 |
| **Moderate maternal anaemia and developmental scores at 5 years** | | | | | | | |
| **Verbal and non-verbal IQ** | 724 | -0.15 (-0.42, 0.12) | 0.28 | 693 | -0.10 (-0.37, 0.18) | | 0.48 |
| **Executive function** | 724 | -0.10 (-0.36, 0.16) | 0.46 | 693 | -0.06 (-0.32, 0.19) | | 0.63 |
| **Motor ability** | 724 | 0.05 (-0.17, 0.27) | 0.64 | 693 | 0.10 (-0.12, 0.33) | | 0.37 |

CI, confidence interval. Mild maternal anaemia was defined as haemoglobin levels <11 g/dL and moderate maternal anaemia was defined as haemoglobin levels <10 g/dL. All haemoglobin measures were adjusted for high altitude (1000 m above sea level). *The multivariable models were adjusted for age at developmental assessment, sex, stunting, socioeconomic status, maternal education, helminthic infections, malaria parasitaemia and any moderate child anaemia event during follow up.

### **Table 10:** Univariable and multivariable linear regression results for the association between measures of iron status and anaemia at two years of age and cognitive and motor scores at five years of age

| **Iron parameter** | **n** | **Univariable model, β (95% CI)** | **P value** | **n** | ***BRINDA adjusted model**  **β (95% CI)** | **P value** |
| --- | --- | --- | --- | --- | --- | --- |
| **Verbal/non-verbal IQ** | | | | | | |
| ID | 490 | 0.11 (-0.14, 0.36) | 0.37 | 413 | 0.22 (-0.05, 0.49) | 0.11 |
| IDA | 472 | -0.08 (-0.42, 0.26) | 0.66 | 422 | 0.02 (-0.24, 0.29) | 0.85 |
| Ferritin (μg/L) | 495 | -0.09 (-0.21, 0.02) | 0.43 | 413 | -0.01 (-0.02, 0.002) | 0.12 |
| Transferrin (g/L) | 517 | 0.17 (-0.03, 0.36) | 0.09 | 431 | 0.12 (-0.09, 0.33) | 0.27 |
| sTfR (mg/L) | 518 | -0.12 (-0.29, 0.04) | 0.15 | 429 | -0.02 (-0.06, 0.01) | 0.15 |
| Hepcidin (μg/L) | 516 | -0.07 (-0.16, 0.03) | 0.16 | 426 | -0.01 (-0.01, 0.003) | 0.22 |
| **Executive Function** | | | | | | |
| ID | 490 | 0.29 (0.05, 0.53) | 0.02 | 413 | 0.06 (-0.20, 0.31) | 0.67 |
| IDA | 472 | -0.08 (-0.41, 0.25) | 0.65 | 422 | -0.05 (-0.30, 0.20) | 0.68 |
| Ferritin (μg/L) | 495 | -0.13 (-0.24, -0.02) | 0.02 | 413 | -0.01 (-0.02, 0.003) | 0.22 |
| Transferrin (g/L) | 517 | 0.22 (0.04, 0.41) | 0.02 | 431 | 0.15 (-0.05, 0.36) | 0.15 |
| sTfR (mg/L) | 518 | -0.03 (-0.19, 0.13) | 0.72 | 429 | -0.01 (-0.04, 0.02) | 0.43 |
| Hepcidin (μg/L) | 516 | -0.12 (-0.21, -0.03) | 0.01 | 426 | -0.01 (-0.01, 0.003) | 0.17 |
| **Motor Ability** | | | | | | |
| ID | 490 | 0.13 (-0.07, 0.33) | 0.21 | 413 | 0.06 (-0.16, 0.29) | 0.57 |
| IDA | 472 | 0.08 (-0.19, 0.35) | 0.58 | 422 | 0.07 (-0.14, 0.29) | 0.50 |
| Ferritin (μg/L) | 495 | -0.08 (-0.17, 0.01) | 0.09 | 413 | -0.01 (-0.02, -0.002) | 0.02 |
| Transferrin (g/L) | 517 | 0.01 (-0.15, 0.16) | 0.91 | 431 | 0.03 (-0.14, 0.21) | 0.71 |
| sTfR(mg/L) | 518 | -0.09 (-0.23, 0.04) | 0.18 | 429 | -0.01 (-0.04, 0.01) | 0.35 |
| Hepcidin (μg/L) | 516 | -0.03 (-0.11, 0.04) | 0.38 | 426 | -0.01 (-0.01, 0.001) | 0.07 |

CI, confidence interval; sTfR, soluble transferrin receptor; ID, iron deficiency; IDA, iron deficiency anaemia. ID was defined as plasma ferritin < 12 μg/L in the absence of inflammation or < 30 μg/L in the presence of inflammation (CRP >5 mg/L); Iron deficiency anaemia was defined as the presence of both iron deficiency and mild anaemia (haemoglobin <11 g/dL). All haemoglobin measures were adjusted for change in altitude (1000 m above sea level). The multivariable models were adjusted for age at iron measurement, sex, stunting, inflammation, helminthic infections, socioeconomic status, maternal education, and malaria parasitaemia. Ferritin, sTfR, hepcidin and CRP levels were natural log (ln) transformed to normalize their distribution. * Iron profiles are regression corrected for inflammation and malaria using the BRINDA approach [26].

### **Figure 1:** Multivariable analyses between annual haemoglobin levels and developmental scores at 5 years

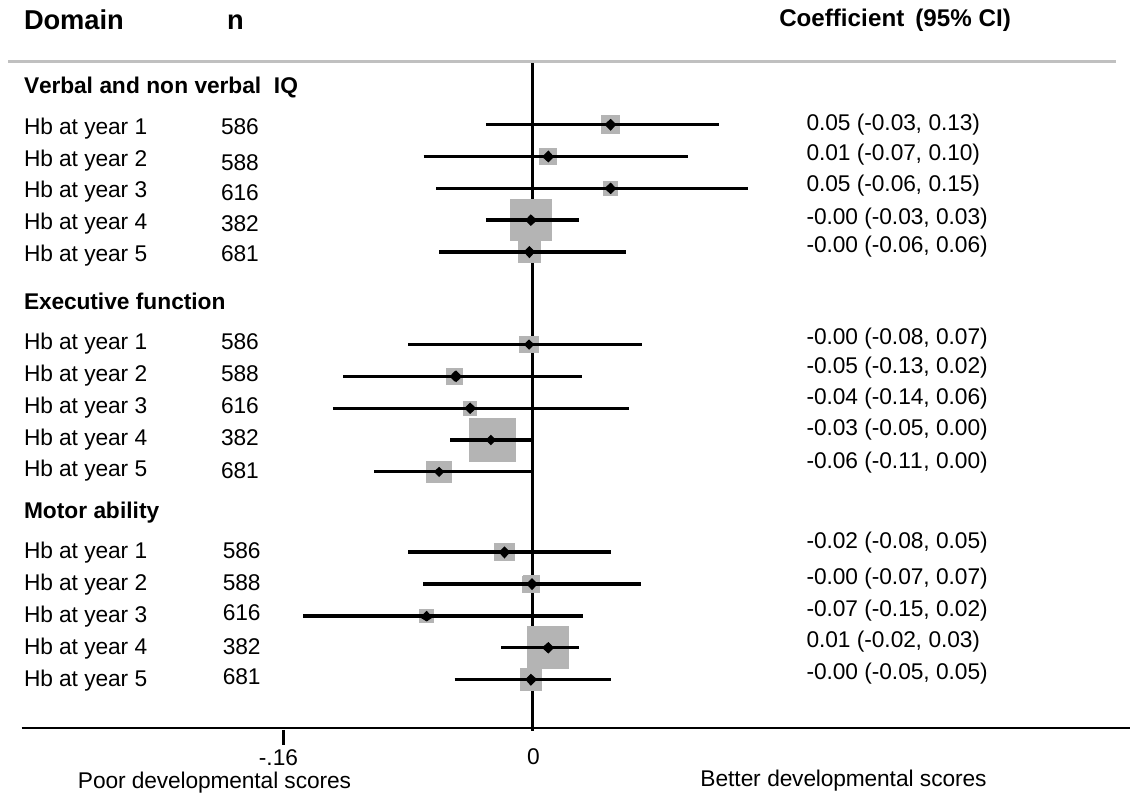

CI, confidence interval; Hb, haemoglobin levels. A forest plot of the multivariable regression analyses between annual haemoglobin levels and developmental scores at five years adjusted for age at developmental assessment, sex, stunting, socioeconomic status, maternal education, helminthic infections and malaria.
